# A facility and community-based assessment of scabies in rural Malawi

**DOI:** 10.1101/2020.09.05.20188557

**Authors:** Cristina Galván-Casas, Oriol Mitjá, Sara Esteban-Terradillos, Jacob Kafulafula, Texon Phiri, Íñigo Navarro-Fernández, Concepción Román-Curto, Hassani Mtenje, Gerald Thauzeni, Elizabeth Harawa, Stephano Kaluzi, Mphatso Diere, Mary Mkandawire, Shaibu Malizani, Alex Chifundo, Marta Utrera-Busquets, Mónica Roncero-Riesco, Sara López Martín-Prieto, Iosune Vilanova-Urdániz, Gisela H. Petiti, María Victoria de Gálvez Aranda, Nuria NO Pérez, María Rueda Gómez-Calcerrada, Pilar Iranzo, Pilar Escalonilla García-Patos, Magdalena de Troya-Martín, Javier Romero Gomez, Esther Cardeñoso-Alvarez, Sofia Lucas Truyols, Libe Aspe Unanue, Cristina Bajo del Pozo, Alicia Comunión Artieda, Maria Isabel Martínez González, Omar F López-López, Esther Moreno-Artero, Xavier Cubiró, Iago Meilán-Sánchez, Alejandra Tomás-Velázquez, Cristina López-Sánchez, Eva M Sánchez-Martínez, Harrison A Edwards, Maria Herrera Morueco, Julia Zehe Rubiera, Laura Salguero Caldera, Urbano Blanes-Moreno, Maria Uribarren-Movilla, Michael Marks

## Abstract

**Background:** Scabies is a neglected tropical disease of the skin, causing severe itching, stigmatizing skin lesions and systemic complications. Since 2015, the DerMalawi project provides an integrated skin diseases clinics and Tele-dermatology care in Malawi. Clinic-based data suggested a progressive increase in scabies cases observed. To better identify and treat individuals with scabies in the region, we shifted from a clinic-based model to a community-based outreach programme.

**Methods:** From May 2015, DerMalawi project provide integrated skin diseases and Tele-dermatological care in the Nkhotakota and Salima health districts in Malawi. Demographic and clinical data of all patients personally attended are recorded. Due to a progressive increase in the number of cases of scabies the project shifted to a community-based outreach programme.

For the community outreach activities, we conducted three visits between 2018 to 2019 and undertook screening in schools and villages of Alinafe Hospital catchment area. Treatment was offered for all the cases and school or household contacts.

**Results:** Scabies increased from 2.9% to 39.2% of all cases seen by the DerMalawi project at clinics between 2015 to 2018. During the community-based activities approximately 50% of the population was assessed in each of three visits. The prevalence of scabies was similar in the first two rounds, 15.4% (2392) at the first visit and 17.2% at the second visit. The prevalence of scabies appeared to be lower (2.4%) at the third visit. The prevalence of impetigo appeared unchanged and was 6.7% at the first visit and 5.2% at the final visit.

**Conclusions:** Prevalence of scabies in our study setting was very high suggesting that scabies is a major public health problem in parts of Malawi. Further work is required to more accurately assess the burden of disease and develop appropriate public health strategies for its control.

**PLAIN ENGLISH SUMMARY:** Scabies is an infestation of the skin caused by a mite. There is limited data on how common scabies is in sub-Saharan Africa, including Malawi. The DerMalawi project has been providing care for dermatological conditions in rural Malawi since 2015. Between 2015 and 2018 we observed an increase in patients with scabies attending for treatment. In response, the project shifted from providing care at clinics to an approach using community-based outreach.

Between 2018 and 2019 we conducted community-based activities on three occasions in an area of approximately 30,000 individuals. The DerMalawi team visited schools and villages to identify and treat cases of scabies and their contacts. We were able to examine about 50% of the population on each visit.

Initially a large proportion of the population had scabies (15%) and this was similar during our second community survey. At our third survey this appeared to have decreased to 2% but it is difficult for us to know if this is because of treatment given in the previous rounds.

Scabies is a major problem in rural populations in Malawi and public health strategies are needed to reduce the disease burden.

## Background

Scabies is a skin disease caused by infestation with the mite, *Sarcoptes scabiei var. Hominis*. Human transmission is predominantly via skin-to-skin contact. Around 200–300 million people are affected each year [1,2], particularly among poor populations living in crowded conditions in tropical areas. There are limited data on the epidemiology of scabies in sub-Saharan Africa and in particular limited population level estimates of disease prevalence.

Permethrin 5% topical ointment is widely considered the first line drug for treatment of individuals with scabies [3] but there is increasing evidence that community wide treatment with ivermectin is the best strategy for control in highly endemic areas. The major challenge to the use of ivermectin is the current recommendation for two separated doses and its relative contraindication in pregnant and breastfeeding women and children under five years of age.[4–9].

Standard guidelines for scabies control are based on treatment of cases and contacts [10], but this approach is not effective in highly-endemic areas [11] [12,13]. Different mass treatment have resulted in positive outcomes [14–16] Mass drug administration of ivermectin as part of lymphatic filariasis control in Zanzibar was approaches associated with a 68% decrease of scabies treatment’s prescription [17]. The first comparative trial of mass drug administration for scabies was the Skin Health Intervention Fiji Trial (SHIFT) which found that ivermectin-based mass drug administration decreased the prevalence of scabies by 94% one year after the intervention, and was superior to both mass drug administration of topical permethrin and standard care [18].

Better data is needed on the epidemiology of scabies in sub-Saharan Africa. We report both facility based and community-based data on scabies in rural Malawi, collected as part of the DerMalawi project.

## Methods

### Ethics Statement

Data reported were routinely collected as part of the activities of the DerMalawi project. The DerMalawi project was established in May 2015 to provide free, integrated skin diseases care and Tele-dermatological consultations to communities in the Nkhotakota and Salima health districts in Malawi. The DerMalawi project is governed by a memorandum of understanding between a group of Spanish dermatologists and the Malawian Health authorities with approval for all activities in the district granted by the Nkhotakota health authorities. Patients give verbal consent when attending the DerMalawi project for an assessment of their skin problems. Approval from the Malawi Medical Council and District Hospital for the DerMalawi project are available in supporting information.

#### Clinic-Based Data Collection

Since the beginning of the DerMalawi project, the demographic and clinical data of all patients personally attended are recorded in spreadsheets, using tablets. Among the clinical data we collected personal and family history, symptoms, dermatological examination, complementary examinations performed, coded diagnosis (ICD-10) and prescribed and administered treatment. This data included basic information on the number of individuals assessed and the number of cases of major common skin conditions including scabies, leprosy, fungal infections, ulcers, albinism and skin cancer. During the period between May 2015 to March 2018, clinic-based data suggested that there had been a progressive increase in the proportion and absolute number of cases of scabies that we observed. In response to this observed increase in scabies cases, the DerMalawi project in collaboration with the Malawi health and educational authorities agreed to shift from a clinic-based model to a community-based outreach programme to try and better identify and treat individuals with scabies in the region.

#### Community Based-Data Collection

The community outreach activities took place in the Alinafe Health Center catchment area, Nkhotakota and Salima districts, Malawi, which has a population of approximately 30,000 individuals. The area has 14 schools, 94 villages and five health facilities. For the purpose of project implementation, the zone was divided into basic units of action, formed by schools and the villages related to each school center. Community dermatology outreach activities were conducted in both schools and communities. Data for the community outreach activities were collected at a community level during the delivery of an active case search and treatment programme in schools and villages of Alinafe Hospital catchment area from July 2018 through April 2019. The programme was conducted in three rounds three and six months apart in July 2018, Oct 2018 and April 2019.

Outreach activities were administered to the population of 14 schools and 94 villages on catch-all basis in a subsection of the catchment area normally covered by the DerMalawi clinic-based service. Clinical teams visited the schools, class by class, and in villages attempted to go house by house trying to reach all individuals present at the school or at home. The teams combined Spanish dermatologists, staff from Alinafe Hospital, trained Chichewa-English translators, community health managers and a local guide. Groups were composed by enough males and females to ensure confidentiality during the skin exam of individuals of both sexes. A week before each round we conducted health awareness in the communities to educate the public to the program’s aims through health educators who disseminated information about scabies at community meetings, school meetings and through public notices, pamphlets and posters in cooperation with village leaders. As with our clinic-based data collection we collected based information on the number of individuals seen, demographics but focused clinical examination on scabies and impetigo in view of the marked increase we had seen at the health facility.

#### Examination

In both clinic and community settings a dermatologist carried out a standardized whole skin examination (including genitalia and buttocks – using tents to ensure). Determination of scabies intensity was based on the number of lesions in categories of mild (≤10), moderate (11 to 49), and severe (≥50).

#### Treatment

At both the clinic and in the community, all individuals diagnosed of scabies were offered one dose of oral ivermectin (200 μg per kilogram of body weight), which was taken under the direct observation of study staff. Ivermectin was replaced with topical permethrin cream in the following participants: children who weighed less than 15 kg, women who were pregnant or breast-feeding. For participants who had scabies at baseline, a second dose of the medication that had been provided at baseline was distributed by study staff 7 at spot-check sites; individuals were advised to attend the clinic. We attempted to provide treatment to all class-mates where cases of scabies were found during school visits and to the household of cases of found during either school or household visits.

#### Analysis

For clinic-based data we report the absolute number of cases of scabies and the proportion of all skin problems seen that were due to scabies. For the community data we report the prevalence of scabies (with and without secondary bacterial infection), for each school area, and the overall intervention area.

### Results

The population living in the catchment area of the Alinafe Health Centre is estimated at 29,780 people (2018 census). Between May 2015 and March 2018, the DerMalawi Project conducted 4 health centre-based clinics at which a total of 4,769 people were seen. The number of people seen increased at each skin visit over the period. The proportion and absolute number of scabies diagnosed increased with 13 (2.8%) cases in 2015, 9 (1,9%) in 2016, 146 (14.5%) in 2017 and 792 (38.7%) in 2018 but there were not significant changes in the proportion of cases of other key conditions such as leprosy, impetigo, tumours or fungal infections (Table 1).

**Table 1:** Cases seen during clinic-based activities.

| **Diagnosis** | **2015** | **2016** | **2017** | **2018** |
| --- | --- | --- | --- | --- |
| **Scabies** | 13 (2.9%) | 9 (1.2%) | 146 (15.2%) | 792 (39.2%) |
| **Leprosy** | 5 (1.1%) | 6 (0.8%) | 10 (1.0%) | 4 (0.2%) |
| **Impetigo** | 18 (3.9%) | 24 (3.1%) | 27 (2.8%) | 46(2.3%) |
| **Skin ulcer** | 3 (0.7%) | 4(0.5%) | 8 (0.8%) | 30 (1.5%) |
| **Tumor** | 16 (3.5%) | 28 (3.6%) | 40 (4.2%) | 33 (1.6%) |
| **Fungal disease** | 192 (42.1%) | 243(31.3%) | 334 (34.8%) | 536 (26.6%) |
| **Total** | 456 | 776 | 960 | 2018 |

Community visits took place between July 2018 and April 2019. We saw 15,290 individuals (51.3% of the population) on our first community visit in July 2018, 16,619 individuals (55.8%) on our second community visit in October 2018, and 12,617 individuals (42.4%) during our third community visit in April 2019 (Table 2).

**Table 2:** Individuals seen during community-based activities.

|  |  | **July 2018**<br>**(N=15487)** | **October 2018**<br>**(N=16619)** | **April 2019**<br>**(N=12617)** |
| --- | --- | --- | --- | --- |
| **Age Group** | 0-4yrs | 2384 (15.4%) | 1894 (11.4%) | 2143 (17.0%) |
|  | 5-9yrs | 3411 (22.0%) | 4242 (25.5%) | 2626 (20.8%) |
|  | 10-14yrs | 3994 (25.8%) | 5490 (33.0%) | 2933 (23.2%) |
|  | 15-24yrs | 2402 (15.5%) | 2259 (13.6%) | 2033 (16.1%) |
|  | 24-34yrs | 1344 (8.7%) | 1106 (6.7%) | 1120 (8.9%) |
|  | >=35yrs | 1952 (12.6%) | 1628 (9.8%) | 1762 (14.0%) |
| **Gender** | Female | 8714 (56.3%) | 9152 (55.1%) | 7246 (57.4%) |
|  | Male | 6773 (43.7%) | 7467 (44.9%) | 5371 (42.6%) |
| **Scabies** | Absent | 13095 (84.6%) | 13764 (82.8%) | 12314 (97.6%) |
|  | Present | 2392 (15.4%) | 2855 (17.2%) | 303 (2.4%) |
| **Impetigo** | Absent | 14455 (93.3%) | 15915 (95.8%) | 11959 (94.8%) |
|  | Present | 1032 (6.7%) | 704 (4.2%) | 658 (5.2%) |

The prevalence of scabies was similar during our first two community visits; 15.4% (2392) in July 2018 and 17.2% (2855) in October 2018 but only 2.4% in April 2019. All individuals diagnosed with scabies received treatment with IVM but only 47% of cases seen in the first community visit, and only 55% of cases seen in during the second community visit were seen for a second dose of treatment. As well as index cases, 5,204 individuals and 7,888 individuals received treatment as contacts during these visits. Prevalence of impetigo was 6.7% (1032), 4.2% (704), and 5.2% (658) across the three visits. At each community visit the prevalence of scabies varied markedly between villages (range 5–40%, Figure 1). The prevalence of scabies was highest amongst children under 5 (Table 3) at each visit.

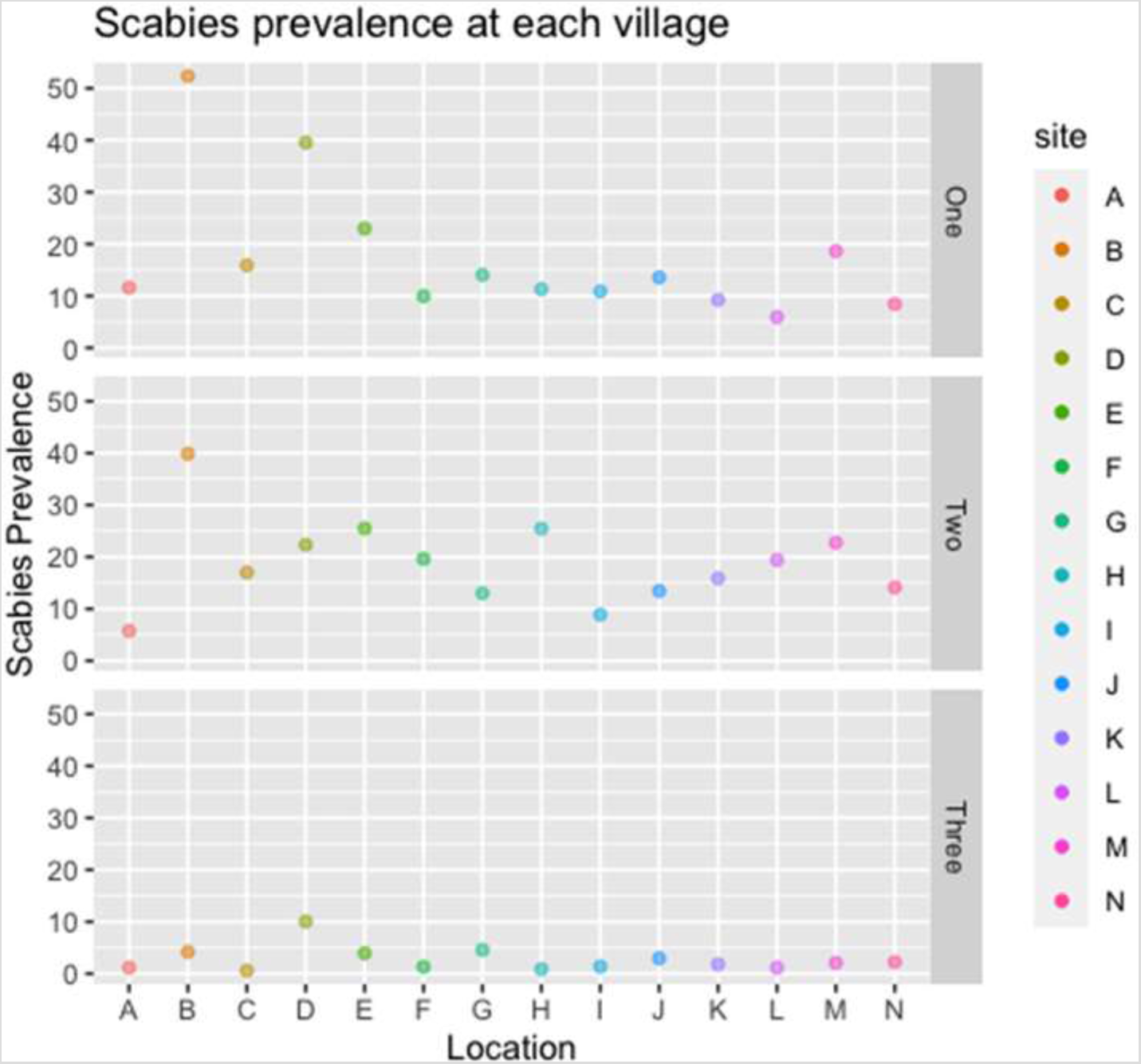
Figure 1

**Table 3:** Prevalence of scabies and impetigo by age group during each visit.

|  | Visit 1 |  | Visit 2 |  | VISIT 3 |  |
| --- | --- | --- | --- | --- | --- | --- |
| Age Group | Prevalence of Scabies | Prevalence of Impetigo | Prevalence of Scabies | Prevalence of Impetigo | Prevalence of Scabies | Prevalence of Impetigo |
| 0-4 | 20.6% | 10.5% | 22.3% | 5.6% | 4% | 6.1% |
| 5-9 | 15.9% | 7.9% | 18.2% | 5.2% | 3.2% | 9.2% |
| 10-14 | 15.8% | 7.2% | 18.7% | 4.7% | 2.8% | 7.1% |
| 15-24 | 14.8% | 5.2% | 15.3% | 3.3% | 1.3% | 2.9% |
| 24-34 | 13.8% | 4.0% | 11.3% | 1.8% | 1% | 1.1% |
| ≥35 | 9.6% | 10.5% | 10.4% | 1.9% | 0.9% | 0.8% |

### Discussion

The DerMalawi project has been working in Malawi since 2015. Over that time we noted a marked increase in presentations to our service for scabies. When we switched to a community rather than a clinic-based model, we found an extremely high community prevalence of scabies of 15%. Over the course of three community outreach activities the prevalence appeared to markedly decline, but whether this reduction is directly related to our activities and treatment or not, remains unclear.

There is limited data on the prevalence of scabies in much of sub-Saharan Africa although several recent studies suggest that it is common [19] and no recent data on scabies in Malawi. The data from the DerMalawi project suggests that scabies is a major problem in the area. We found especially high prevalences amongst children [20–22]

There was no change in scabies prevalence between July 2018 and October 2018 suggesting that at the coverage achieved the treatment of cases and a proportion of their contacts was inadequate to affect scabies prevalence. This lack of change is likely due to low population coverage. Whilst we switched from a clinic to a community-based strategy, we still reached only about 50% of the population of our catchment area. Such low coverage invariably leads to a large number of untreated scabies cases that might act as a reservoir for infection. In addition, most individuals received only a single dose of ivermectin limiting the efficacy of treatment for individual patients. All three visits were conducted during the dry season, so differences in prevalence seem unlikely to be related to seasonality [23]. Unlike research interventions that have been carried out in islands or confined populations and have maintained the effects for years [13], the activities reported here were carried out as part of a long term dermatological assistance programme and the patients in our target area, are surrounded by neighbouring communities with potentially high scabies rates. These surrounding areas therefore serve as a further reservoir from which reintroduction can occur.

The markedly reduced prevalence observed in our visit in April 2019 despite low treatment coverage, and in the absence of any effect seen previously, suggests that the observed reduction is unlikely to be due to treatment and more likely reflects changes in the population assessed at each time point. In keeping with this there were differences in both the demographic structure of the population seen in our final community visit and in the number of people seen at each screening site across each of our visits. We noted that in the final visit fewer people were able to enter the study because they said they were not affected and refused to be examined. What they find most difficult is getting undressed, and they refused to be examined arguing that they did not notice any itching or skin lesions. For these reasons it seems likely that the reduction seen is most likely a reflection of sampling bias not a true difference.

Scabies remains as an unresolved matter for global health, continues to be widespread and is responsible for intense suffering, health and social complications. New drugs, dissemination models and treatment and preventive strategies are needed to achieve better results. Targets for the establishment of scabies control programmes have been included in the new WHO NTD Roadmap 2021–2030 and our data suggests such a programme should be considered in Malawi alongside other national NTD programmes.

## Data Availability

All data are fully available without restriction. All relevant data are within the manuscript and its Supporting Information files.

## Acknowledgments

We are extremely grateful to the Missionary Community San Paul the apostle MCSPA for providing accommodation and support for the DerMalawi project, especially Mr. Brian Odhiambo who supports all the dermatological work carried out by the DerMalawi project and F. Manuel Hernandez, head of the parish. We are also extremely grateful to all the members of the community and community leaders for their participation and support of the DerMalawi project over many years.

## Notes

### Competing Interest Statement

The authors have declared no competing interest.

### Clinical Trial

This work is not a Clinical Trial. Data reported were routinely collected as part of the activities of the DerMalawi project. The DerMalawi project was established in May 2015 to provide free, integrated skin diseases care and Tele-dermatological consultations to communities in the Nkhotakota and Salima health districts in Malawi. The DerMalawi project is governed by a memorandum of understanding between a group of Spanish dermatologists and the Malawian Health authorities with approval for all activities in the district granted by the Nkhotakota health authorities. Patients give verbal consent when attending the DerMalawi project for an assessment of their skin problems. Approval from the Malawi Medical Council and District Hospital for the DerMalawi project are available in supporting information.

### Funding Statement

This work was supported by donations from individuals, companies (CantabriaLabs https://www.cantabrialabs.es/ , Martiderm https://www.martiderm.es/ ,Pierre Fabre https://www.pierre-fabre.com/es-es and Italfarmaco https://www.italfarmaco.es/), Ayuntamiento de Villanueva de la Canada https://www.ayto-villacanada.es/ , and students from Secondary School Las Encinas, from Villanueva de la Canada, Madrid (Spain) https://www.ieslasencinas.es/ , Celia Delgado Matias Association http://www.asociacionceliadelgadomatias.org/ http://www.asociacionceliadelgadomatias.org/proyectos/2018-2/ , University of Salamanca https://www.usal.es/ , the collaboration agreement between the Spanish Academy of Dermatology (AEDV) and the The State Foundation, Health, Childhood and Social Welfare, P.S.F. of the Spanish Ministry of Health (former Spanish Foundation for International Cooperation, Health and Social Policy). https://contrataciondelestado.es/wps/poc?uri=deeplink%3AperfilContratante&idBp=4lhzg%2BRWZr4%3D, the international Cooperation Awards from the Spanish Academy of Dermatology (AEDV) https://aedv.es/ ; https://fundacionpielsana.es/dermatologia-solidaria/la-solidaridad-de-los-dermatologos-tiene-premio , the Colegio Oficial de Medicos de Madrid (ICOMEM) https://www.icomem.es/ https://www.icomem.es/comunicacion/noticias/3343/El-Colegio-de-Medicos-de-Madrid-incrementa-el-numero-de-proyectos-y-la-cuantia-economica-dedicada-a-proyectos-de-cooperacion-internacional-en-salud , the Fundacion Mutua Madrilena (FMM) https://www.fundacionmutua.es/ ; https://www.fundacionmutua.es/FundMM/action/main?execution=e1s1 , and the International Cooperation Award from the Central section of the Spanish Academy of Dermatology (SC-AEDV) https://seccion-centro.aedv.es/ https://seccion-centro.aedv.es/dermatologia-solidaria/ ;
All donations were delivered through Zikomo Africa Organization http://www.zikomoafrica.org/ and Emalaikat Foundation https://fundacionemalaikat.es/

